# The association of *CREBRF* variant rs373863828 with body composition in adult Samoans

**DOI:** 10.1101/2021.02.11.21251582

**Authors:** Nicola L. Hawley, Rachel L. Duckham, Jenna C. Carlson, Take Naseri, Muagututia Sefuiva Reupena, Viali Lameko, Alysa Pomer, Abigail Wetzel, Melania Selu, Vaimoana Lupematisila, Folla Unasa, Lupesina Vesi, Tracy Fatu, Seipepa Unasa, Kima Faasalele-Savusa, Satupaitea Viali, Anna C. Rivara, Emily M. Russell, Ranjan Deka, Erin E. Kershaw, Ryan L. Minster, Daniel E. Weeks, Stephen T. McGarvey

## Abstract

**Objective:** The minor allele of rs373863828, a missense variant in *CREBRF*, is associated with higher BMI but lower odds of type 2 diabetes in Pacific Islanders.

**Methods:** To test if this protective effect operates through metabolically favorable body fat distribution, we examined the association of the minor A allele with body composition, measured using dual-energy x-ray absorptiometry (DXA), in a cross-sectional study of *n*=421 Samoan adults.

**Results:** We replicated our earlier finding that this allele was associated with higher weight and BMI, although it was statistically significant only in women. There was no significant association of genotype with percent body fat, visceral adiposity or fat distribution, although in women, the A allele was associated with greater total fat mass (*p*=0.02), android (*p*=0.009) and trunk fat (*p*=0.01). In both sexes, age- and height-adjusted average lean mass was significantly greater per copy of the A allele: 2.16 kg/copy in women and 1.73 kg/copy in men.

**Conclusions:** These data do not support a primary role of fat distribution in mediating the association between rs373863828 genotype and type 2 diabetes risk. We suggest an alternative hypothesis: those with the A allele may more efficiently regulate blood glucose because of their greater absolute lean mass.

## Introduction

In 2016, based on a genome-wide association study (GWAS) of body mass index (BMI) in adult Samoans, we reported the identification of a missense variant in CREB3 regulatory factor (*CREBRF*) that is paradoxically associated with higher BMI and odds of obesity but with lower fasting blood glucose and odds of type 2 diabetes [1]. The minor allele A of rs373863828, an arginine-to-glutamine missense variant, had a frequency of 0.259 in the discovery sample, indicating that >40% of Samoans have at least one copy of the risk allele.

Our discovery has been replicated in several other Pacific Islander populations (Māori, Tongans, Cook Islanders, Niueans, Chamorro, Chuukese) [2-6] and may partially explain their greater risk of obesity compared to other ethnic groups. The paradoxical associations of rs373863828 with BMI and diabetes, however, remains unexplained. In a murine 3T3L1 adipocyte model, ectopic expression of the human variant enhanced adipogenesis and lipid storage compared to control [1]. Therefore, we hypothesized that human carriers of the variant would have greater fat mass relative to lean mass but would store that fat in a more metabolically favorable distribution (e.g., subcutaneously rather than viscerally, or peripherally rather than centrally) than those without the variant, thereby explaining their lower odds of diabetes.

To test that hypothesis, we examined the association between rs373863828 genotype and body composition, measured using dual-energy x-ray absorptiometry (DXA).

## Methods

Between August 2017 and March 2019 participants from the original GWAS sample were recruited into a follow-up study to examine body composition and cardiometabolic health. Participants with AA, AG, and GG genotypes were targeted in a 1:2:2 ratio. Protocols for the original GWAS in 2010 and the follow-up study, including genotyping methods, have been previously published [7,8].

Eligible participants were not pregnant, lactating, or attempting to control their weight through medication or surgery, were resident on the island of ‘Upolu and were part of a maximally unrelated sample [maximum kinship 6.01%] [8]. Willing participants (*n*=519; aged 30.7-72.7 years) gave written informed consent. Protocols were approved by the Yale University Institutional Review Board (IRB #1604017547), the University of Pittsburgh (#PRO16040077), and the Health Research Committee of the Samoan Ministry of Health.

Weight and height were measured using a Tanita HD 351 digital weighing scale (Tanita Corporation of America, IL) and SECA 213 portable stadiometer (Seca GmbH & Co., Germany), respectively. DXA outcomes (assessed using a Lunar iDXA, GE Healthcare Medicine, Encore Version 17) included total body fat, lean, and bone mass. Visceral fat mass (estimated using GE CoreScan ™) was also measured along with fat mass in the android (central) and gynoid (hip and thigh) regions and the limbs. Android-to-gynoid fat ratio (a measure of abdominal vs. gluteal fat distribution) was calculated, as well as trunk-to-peripheral fat ratio (TPFR), a measure of centrality of fat deposition (fat mass in the trunk region divided by the sum of arm and leg fat [peripheral fat]). When participant’s width exceeded the scan area, a right-side scan was ‘mirrored’ and used to estimate total body composition [9]. Contraindications to DXA included exposure to additional X-rays or computed tomography (CT) in the prior 12 months. Therefore, this analysis was restricted to *n*=421 participants (AA *n*=72, AG *n*=161, GG *n*=188). For analyses of visceral fat mass, the sample was further reduced by *n*=1 participant whose body size prevented the capture of the core region in a single scan.

Associations between body size and composition outcomes and rs373863828 genotype were performed using linear models that were sex-stratified and adjusted for age and age^2^ (sex and age were self-reported by participants). Body composition outcomes were also adjusted for height [10]. Genotype was modeled additively (the number of A alleles a participant carries), consistent with previous work [1]. Analyses were conducted in R Version 3.6.0 (R Foundation for Statistical Computing, Austria) with the threshold for statistical significance set at *p*<0.05. For ease of interpretation, effect sizes are presented on their original scale; sensitivity analyses using inverse-normally transformed traits gave similar results.

## Results

The rs373863828 minor A allele was associated with higher average weight and BMI, significantly so among women, but not men (**Table 1**). Average total fat mass (*p*=0.02), android (*p*=0.009), and trunk fat mass (*p*=0.01) were significantly higher with each copy of the minor allele among women but not men (**Table 1, Figure 1**), although the same trend toward higher fat mass was present in men. In both sexes, average lean mass was greater with each copy of the minor allele: by 2.16 kg/copy in women (*p*=0.0001) and 1.73 kg/copy in men (*p*=0.02) after a6-7 (tdjusting for age, age^2^, and height. There were no significant differences by genotype observed among either sex in visceral adiposity, percent body fat, nor in the distribution of body fat based on android-to-gynoid ratio and TPFR (**Table 1**). Additional plots describing distribution of body size and composition outcomes by genotype are provided in **Supplementary Figure 1**.

**Table 1:** Age, body size, and body composition by genotype.

|  | Female |  |  |  |  | Male |  |  |  |  |
| --- | --- | --- | --- | --- | --- | --- | --- | --- | --- | --- |
| | GG<br>(n=103) | AG<br>(n=92) | AA<br>(n=38) | $\beta$ | *p* | GG<br>(n=85) | AG<br>(n=69) | AA<br>(n=34) | $\beta$ | *p* |
| Age (years) | 50.8 (9.5) | 50.6 (9.8) | 50.0 (9.0) | -0.36 | 0.67 | 53.1 (10.3) | 53.7 (9.3) | 49.0 (11.0) | -1.59 | 0.11 |
| Body Mass Index (kg/m<sup>2</sup>)<sup>a</sup> | 35.8 (6.4) | 37.1 (6.2) | 40.3 (9.2) | 1.95 | 0.0014 | 32.7 (5.7) | 33.2 (5.7) | 34.0 (5.9) | 0.60 | 0.32 |
| Weight (kg)<sup>a</sup> | 94.2 (19.3) | 97.3 (17.5) | 106.5 (24.9) | 5.21 | 0.002 | 95.7 (21.2) | 99.4 (18.1) | 103.8 (19.8) | 3.67 | 0.06 |
| Height (cm)<sup>a</sup> | 161.8 (5.6) | 161.8 (5.3) | 162.6 (6.6) | 0.24 | 0.62 | 170.9 (6.4) | 173.0 (6.0) | 174.8 (5.5) | 1.71 | 0.003 |
| Total Fat Mass (kg)<sup>b</sup> | 40.9 (12.6) | 42.6 (11.1) | 47.0 (15.8) | 2.51 | 0.02 | 29.1 (12.7) | 29.8 (11.7) | 30.3 (13.0) | -0.11 | 0.93 |
| Total Lean Mass (kg)<sup>b</sup> | 50.3 (7.4) | 51.7 (7.5) | 55.7 (8.3) | 2.16 | 0.0001 | 63.2 (9.4) | 66.2 (8.2) | 70.1 (8.0) | 1.73 | 0.02 |
| Total Bone Mass (kg)<sup>b</sup> | 2.5 (0.4) | 2.5 (0.4) | 2.7 (0.4) | 0.03 | 0.23 | 3.3 (0.5) | 3.4 (0.4) | 3.5 (0.4) | 0.02 | 0.51 |
| Visceral Fat Mass (kg)<sup>b</sup> | 1.33 (0.6) | 1.47 (0.6) | 1.49 (0.6) | 0.09 | 0.10 | 1.9 (1.1) | 2.0 (1.1) | 1.8 (1.1) | -0.01 | 0.96 |
| Missing | 0 | 0 | 0 |  |  | 0 | 1 | 0 |  |  |
| Percent Fat (%)<sup>b</sup> | 42.8 (6.0) | 43.5 (5.0) | 43.6 (5.7) | 0.46 | 0.36 | 29.2 (7.3) | 29.1 (7.1) | 28.1 (7.5) | -0.38 | 0.60 |
| Android Fat Mass (kg)<sup>b</sup> | 3.7 (1.3) | 3.9 (1.2) | 4.4 (1.6) | 0.31 | 0.009 | 3.0 (1.6) | 3.1 (1.5) | 3.0 (1.6) | -0.03 | 0.83 |
| Gynoid Fat Mass (kg)<sup>b</sup> | 6.1 (2.1) | 6.3 (1.8) | 6.9 (2.7) | 0.30 | 0.09 | 4.2 (1.9) | 4.2 (1.8) | 4.4 (1.9) | -0.07 | 0.70 |
| Android:Gynoid Fat Ratio<sup>b</sup> | 0.6 (0.1) | 0.6 (0.1) | 0.7 (0.1) | 0.02 | 0.07 | 0.7 (0.2) | 0.7 (0.2) | 0.7 (0.2) | 0.02 | 0.34 |
| Trunk Fat Mass (kg)<sup>b</sup> | 21.7 (6.6) | 22.8 (6.0) | 25.1 (8.3) | 1.50 | 0.01 | 16.8 (8.0) | 17.5 (7.1) | 17.1 (7.7) | -0.11 | 0.88 |
| Peripheral Fat Mass (kg)<sup>b,c</sup> | 18.2 (6.3) | 18.7 (5.4) | 20.7 (7.9) | 0.97 | 0.07 | 11.2 (4.8) | 11.2 (4.7) | 12.0 (5.3) | -0.02 | 0.97 |
| Trunk-to-Peripheral Fat Ratio<sup>b</sup> | 1.2 (0.2) | 1.2 (0.2) | 1.3 (0.3) | 0.02 | 0.34 | 1.5 (0.3) | 1.6 (0.3) | 1.4 (0.3) | 0.01 | 0.62 |

Values are means (standard deviation). Beta values are the estimated difference in mean per copy of the minor (A) allele (between AA and AG or AG and GG) from a linear model.
<sup>a</sup>Linear models were adjusted for age and age<sup>2</sup>
<sup>b</sup>Linear models were adjusted for age, age<sup>2</sup>, and height
<sup>c</sup>Peripheral fat mass was calculated from the sum of limb (arm and leg) mass

**Figure 1:**
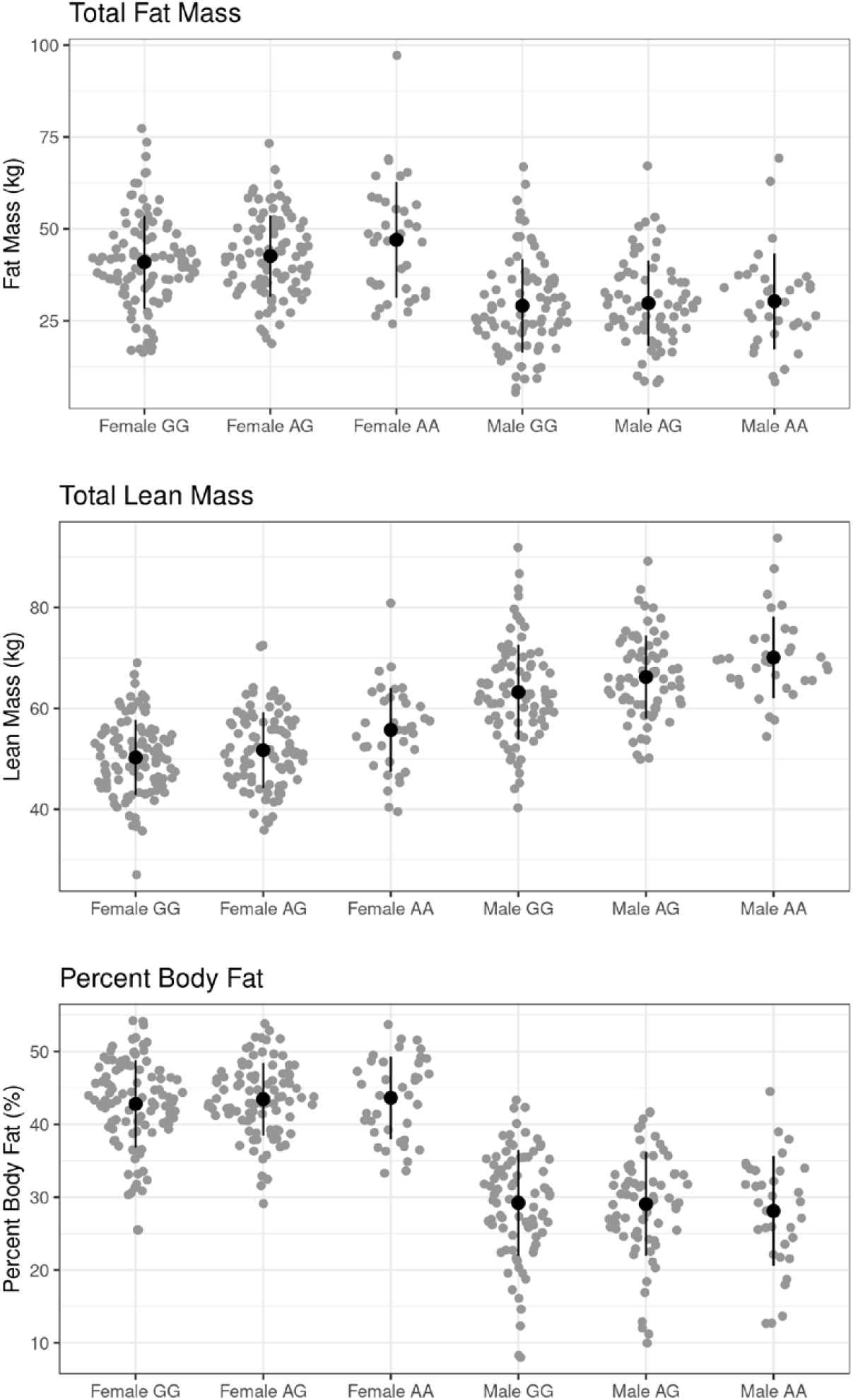
Fat, lean, and bone mass and percent body fat by genotype.

## Discussion

In this study we demonstrate that the original association between the rs373863828 minor A allele and BMI persisted after 8 years. Moreover, we observed that the positive effect of the allele on BMI is primarily a function of greater lean mass rather than fat mass among men, and a combination of greater lean and fat mass among women. The findings do not support the hypothesis that the inverse association between the A allele and type 2 diabetes is mediated by more metabolically favorable fat distribution. They are, however, consistent with our recent work in infants where we observed greater lean mass at four months of age [15] among those with the A allele. While it is not possible to obtain the same measures of fat distribution measures in infants, we saw no effect on total or percent fat mass.

An alternative hypothesis, consistent with these data, is that those with the A allele may more effectively regulate blood glucose as a result of their greater lean mass. Greater muscle and bone mass promote lower serum glucose, greater insulin sensitivity, and lower risk of diabetes via multiple mechanisms [11,12]. Skeletal muscle, in particular, is a major site for insulin-dependent and independent glucose uptake and disposal as well as secretion of autocrine, paracrine, and endocrine factors that influence metabolic homeostasis, such as irisin, interleukin-6, and other myokines [13,14]. While outside the scope of this report, we will test this hypothesis in future work by using structural equation modeling to examine the effect of rs373863828 on fasting glucose and the odds of type 2 diabetes as mediated through its direct and indirect effects on lean and fat mass. Further research, using CT or magnetic resonance imaging (MRI) to distinguish between muscle and other lean tissue, which DXA cannot do, will be needed to fully test this new hypothesis.

While the minor allele was associated with greater lean mass in both men and women, and height in men (consistent with our earlier work [16]), we observed a stronger effect of the missense variant on BMI in women and an effect on fat mass in women only. Prior studies of this variant and its association with BMI have adjusted for sex, rather than presenting sex-stratified analyses (likely because of their limited sample size) [2-6], so this phenomenon has not been noted previously. Our findings indicate that attempts to examine potential sex-specific effects of the variant are warranted.

The major strength of this study compared to other examinations of the rs373863828 genotype to date is the use of DXA-measured body composition, rather than BMI, which is unable to distinguish between fat and lean mass. We did, however, encounter many participants (19% of our recruited sample) who, because of our conservative safety criteria, could not participate due to recent high dose radiation exposure. While those who could not receive a DXA scan were similar in age, BMI, and genotype distribution, this may limit generalizability of our findings. Furthermore, we note that the participants in our study sample had high average levels of body fat (approximately 30% in men and 45% in women). Future studies are needed to explore the functional relationship between fat distribution and rs373863828 across the spectrum of adiposity, including examining the impact upon cardiometabolic disease risk factors.

With further study of insulin-glucose metabolism and its association with the rs373863828 variant ongoing among several Pacific Islander groups, these findings indicate that greater lean mass among those with the minor allele may be responsible for the partially protective effect of the minor A allele on odds of diabetes. As diabetes prevalence continues to increase among Samoans and other Pacific Islander populations, additional understanding of the mechanisms that underlie this association may be transformative for prevention and treatment.

## Supporting information

STREGA checklist

## Data Availability

The data that support the findings of this study are available on request from the corresponding author NJH.

