## Supplementary Figure 1 for "The association of *CREBRF* variant rs373863828 with body composition in adult Samoans"

**Supplementary Figure 1: Distribution of additional body size and body composition outcomes by genotype**

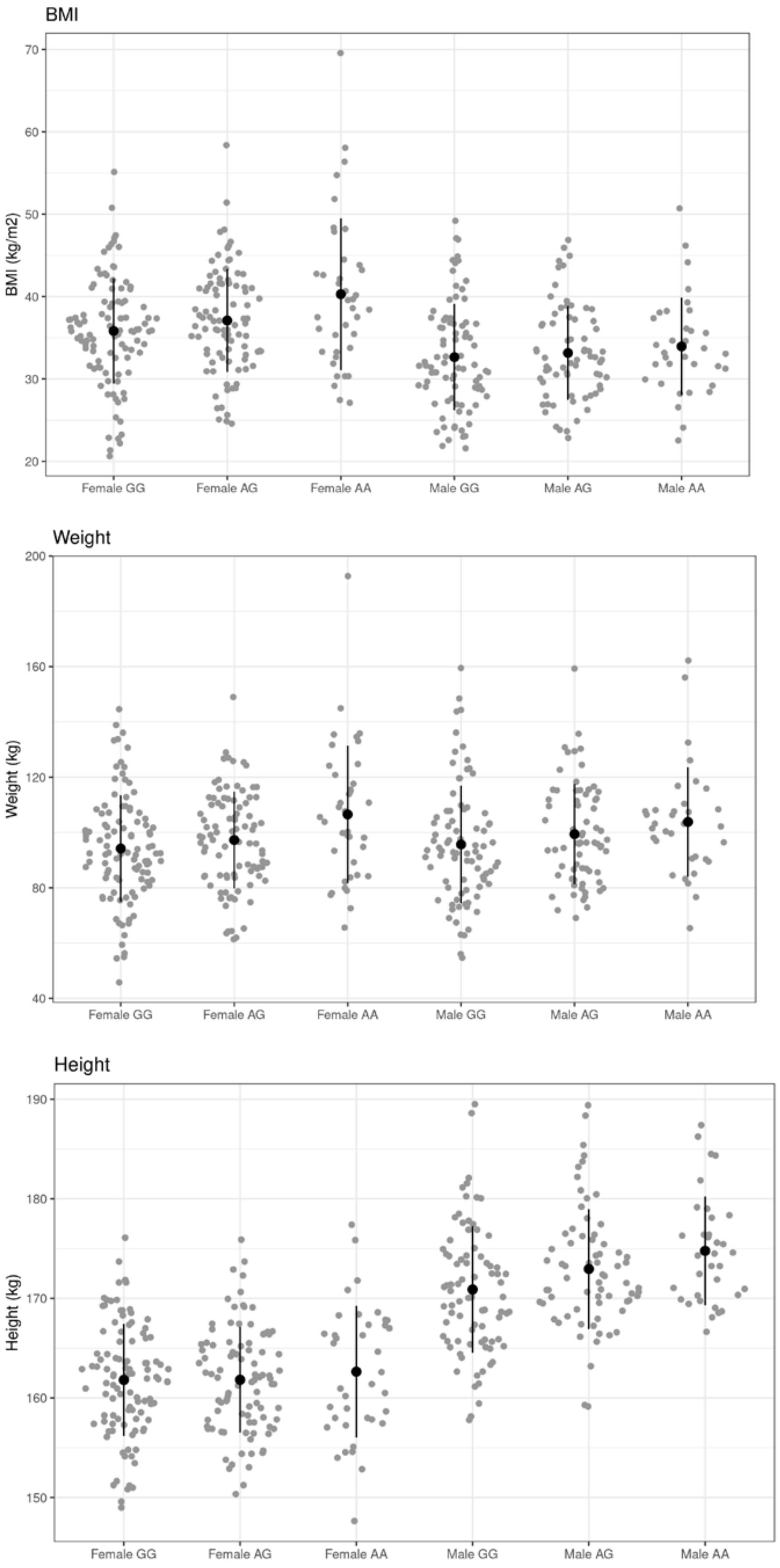

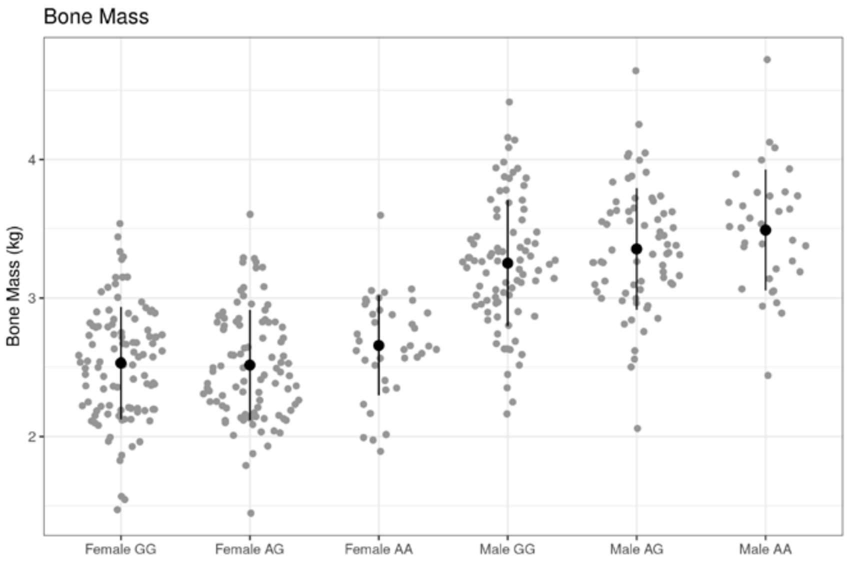
Bone Mass
Bone Mass (kg)
2
3
4
Female GG
Female AG
Female AA
Male GG
Male AG
Male AA

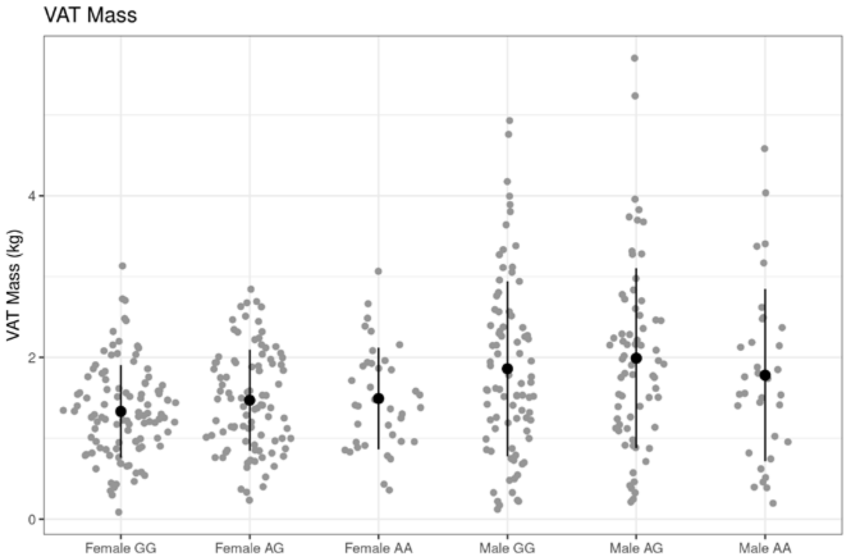
VAT Mass
VAT Mass (kg)
0
2
4
Female GG
Female AG
Female AA
Male GG
Male AG
Male AA

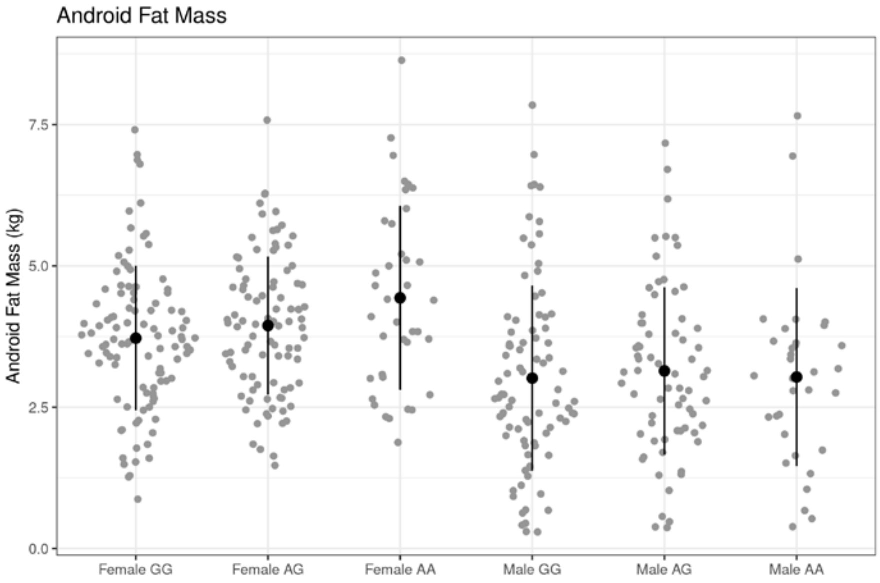
Android Fat Mass
Android Fat Mass (kg)
0.0
2.5
5.0
7.5
Female GG
Female AG
Female AA
Male GG
Male AG
Male AA

Gynoid Fat Mass

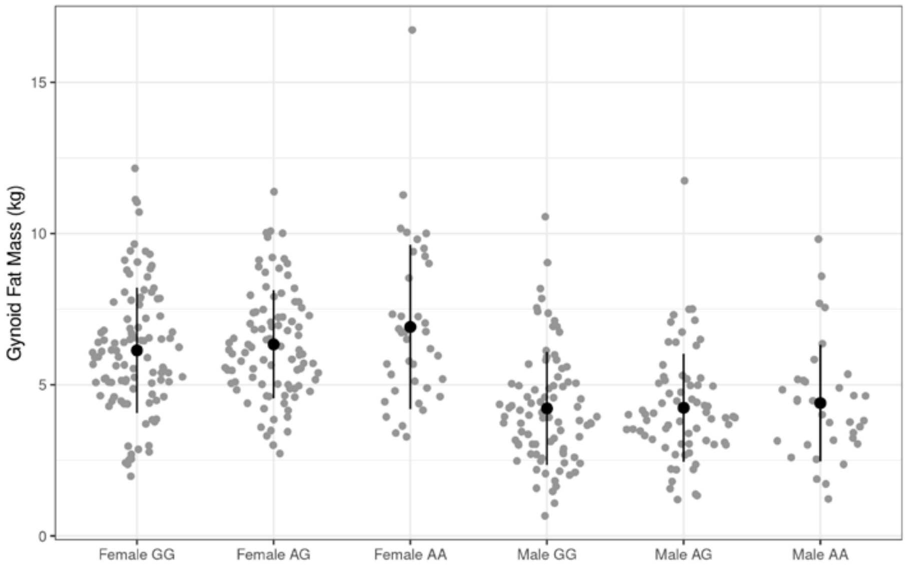


Anroid Gynoid Ratio

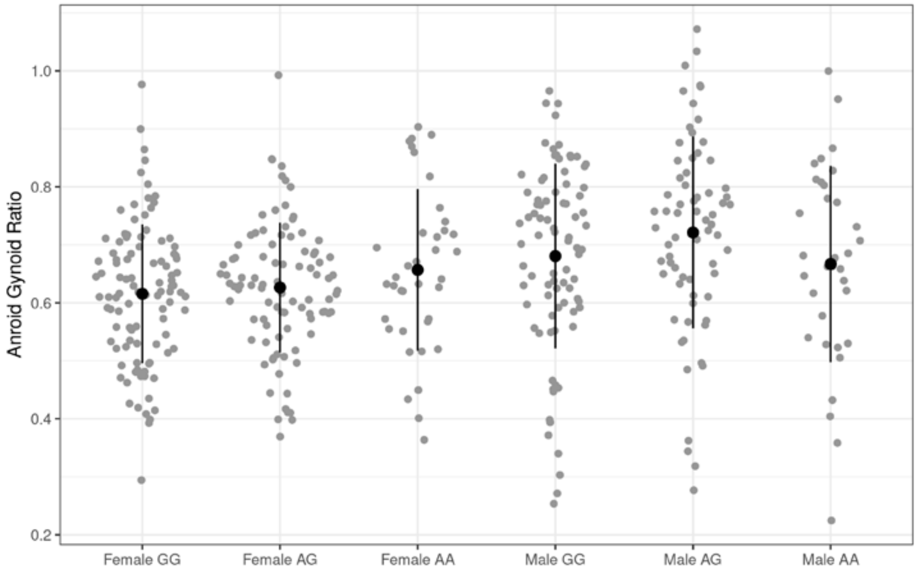


Trunk Fat Mass

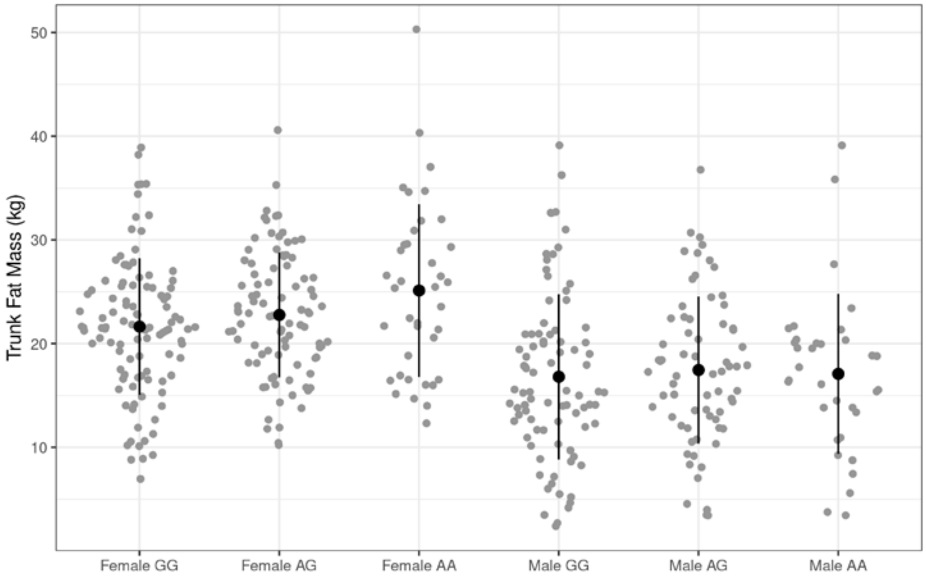

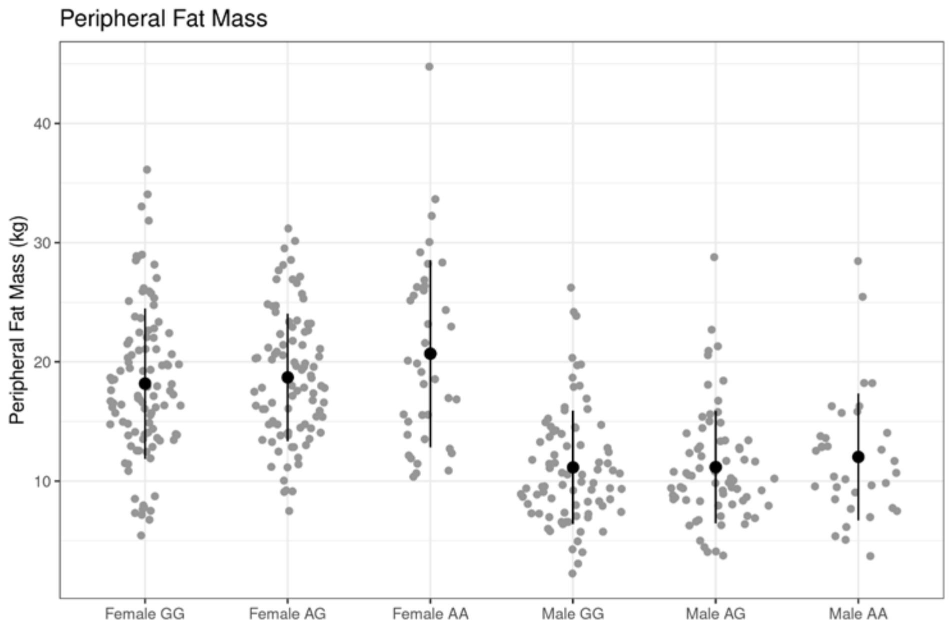


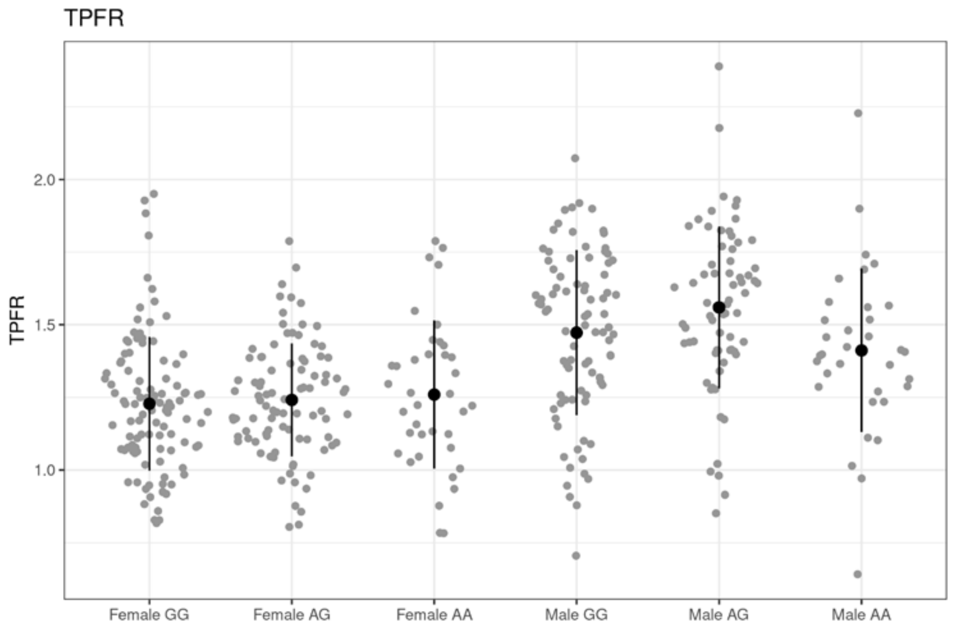


Abbrevations: BMI: Body Mass Index; VAT: Visceral Adipose Tissue; TPFR: Trunk to peripheral fat ratio
